# Invisible Spectrum: A model for minority community public engagement in cancer research

**DOI:** 10.1101/2024.12.02.24317617

**Authors:** Shane O’Grady, Jessica C. Ralston, Eadaoin McKiernan, Fiona Lanigan, Md Salauddin, Shayla Sharmin, Terri McVeigh, William M. Gallagher, Suzanne Guerin, Arman Rahman, Walter Kolch

## Abstract

**Background:** Ethnic minority communities are often recognised as experiencing decreased accessibility to vital medical services as well as increased barriers to participation in research studies. These issues stem from a variety of social, cultural and economic factors, all of which must be taken into consideration when designing engagement initiatives for a particular community. *Invisible Spectrum* is an annual engagement initiative which seeks to promote effective communication and outreach to often-overlooked ethnic minority communities within Ireland, primarily those of Bangladeshi origin. The programme was developed in response to the traditionally low levels of engagement with healthcare services observed within these communities and seeks to empower communities in their own healthcare decision making.

**Methods:** This study reports a programme of community participation activity with an embedded empirical research component based on participatory action research. The team included researchers and leaders within the Bangladeshi and Arabic Muslim communities in Ireland. Over the course of four years, feedback polls, pre- and post-event surveys and in-depth interviews gathered the views and recommendations of attendees.

**Results:** Four annual events were held as part of the *Invisible Spectrum* programme, from 2019 to 2023. Feedback from participants consistently demonstrated high levels of satisfaction within the target communities while quantitative survey data also indicated improvements in key areas such as recognition of potential cancer symptoms and greater awareness of available screening services.

**Conclusion:** There is a significant need to continually promote patient involvement and minority inclusion in healthcare and research initiatives. In line with this goal and after 4 successful years of running the *Invisible Spectrum* programme, we have developed a set of recommendations and guidelines for the successful development and organisation of minority community engagement initiatives. It is our hope that the *Invisible Spectrum* programme could be used as a model for future endeavours of a similar nature.

## Introduction

The relationship between one’s ethnicity and health is a topic of common discussion in healthcare research. This relationship is complex and heterogenous: within cancer, some studies have indicated that within Western nations, minority populations often have increased rates of infection-associated cancers (e.g. liver, cervical, stomach) but lower incidence of cancers associated with a “Westernised” lifestyle (colorectal, breast, prostate) (1, 2). The rates of cancers associated with Western lifestyle has also been observed to rise over time in migrant populations (3), possibly due to changes in lifestyle factors such as diet and consumption of alcohol and tobacco products.

Additionally, minority communities have also been observed to have low levels of participation in scientific research studies (4) and engagement with researchers and clinicians. Reasons for this historical lack of engagement are multifactorial and include factors stemming from both minority communities (social and cultural, trust, language barriers, economic) and researchers/clinicians (lack of understanding of how to begin engagement efforts, discomfort at bringing up a patient’s ethnic background). Indeed, it can be difficult to even ascertain the true extent of the problem. In Ireland, information regarding ethnicity is often uncollected as part of research surveys: Hannigan et al found that only 14% of national health and social care data collections in Ireland contained data regarding ethnicity (5). Most notably, there was no ethnic data recorded in the official cancer registry. A subsequent study looked at reasons behind the lack of ethnicity data gathering by GPs and found that, although many understood the rationale, they were wary of the potentially sensitive nature of such questions and remained sceptical of a genuine healthcare benefit behind gathering such data (6). Concerns were also raised about the validity of grouping together highly diverse populations based on superficial characteristics. For instance, people of African, African-American and Caribbean descent may all be grouped under the term “Black”, despite extensive differences in cultural, genetic and lifestyle factors. This can lead to a “one size fits all” approach, which is the very opposite of culturally aware patient engagement and could lead to further divides in understanding between neglected populations and the medical/scientific community.

Some communities are more affected by these issues than others. Studies from the UK have shown that Bangladeshi schoolchildren were found to perform below the national average in science (7) and Bangladeshi adults were the most under-represented ethnic group in STEM roles (8). Although there is less data on this topic within an Irish context, it is likely that similar patterns are in effect. There is a real and direct human cost to this disconnect between members of an ethnic minority and the medical/scientific communities. As an example, a large-scale study in the UK found that non-White populations scored significantly lower in their ability to recognise potential signs and symptoms of cancer (9). People of Bangladeshi origin scored amongst the lowest, with an almost 7-fold difference in the odds of recognising a lump as a potential sign of cancer, in comparison to White British people. Bangladeshi women were also found to have, at a statistically significant level, a greater likelihood of encountering barriers to accessing medical care, stressing the importance of looking at medical engagement efforts in an intersectional manner. More recently, the Marmot Report highlighted that health and sociocultural inequalities significantly increased the mortality risk for ethnic Bangladeshi males in the UK, who were 1.9 times more likely to die from COVID-19 compared to their white counterparts (10). This all points to a real need for a focused intervention on carefully targeted populations.

The *Invisible Spectrum* programme was developed in response to these issues. Our goal is to promote effective engagement and communication with minority communities in Ireland who have often been underrepresented in previous efforts. The name “*Invisible Spectrum*” evokes the large segments of electromagnetic spectrum that surrounds us at all times and yet remains invisible due to the limits of our sensory systems. In a similar way, due to the traditional bias of scientific research towards homogenous, predominantly Caucasian populations, how can we ensure that those outside of this narrow slice of the spectrum are sufficiently represented in research, medical outreach and vital engagement with healthcare services? *Invisible Spectrum* has evolved and adapted over its lifespan, from an initial pilot event in 2020, held fully online due to COVID-19 limitations, to a comprehensive in-person programme of research talks, gender-segregated breakout sessions for sensitive discussions, participation from clinicians, researchers and patient involvement organisations and even a child friendly “cell biology workshop”.

After running *Invisible Spectrum* for four consecutive years, we have built a significant level of in-house expertise in the organisation of such programmes and feel the outcomes and learnings could be valuable to other groups interested in coordinating similar initiatives. We envision *Invisible Spectrum* as a flagship example of how to design and organise an ethnic minority outreach programme, which promotes effective engagement with these communities while always centring their perspectives and needs. The aim of this paper is to report the key components of the process, along with the findings of a participatory action research component that evaluated the process.

## Methods

### Research Design

This paper reports on the development and implementation of the *Invisible Spectrum* programme, which included a participatory action research (PAR) design component at its core. PAR as a research design seeks to **“**brings together community members, activists and scholars to co-create knowledge and social change” (11). The programme of work includes a co-design process to develop the programme and an analysis of data collected over four phases of delivery of *Invisible Spectrum*.

Figure 1 contains a general representation of our organisational flow. This workflow was developed and refined over the programmes lifespan and represents accumulated learnings from each annual initiative. The research was approved as a low-risk research ethics application by the UCD Human Research Ethics Committee.

**Figure 1.**
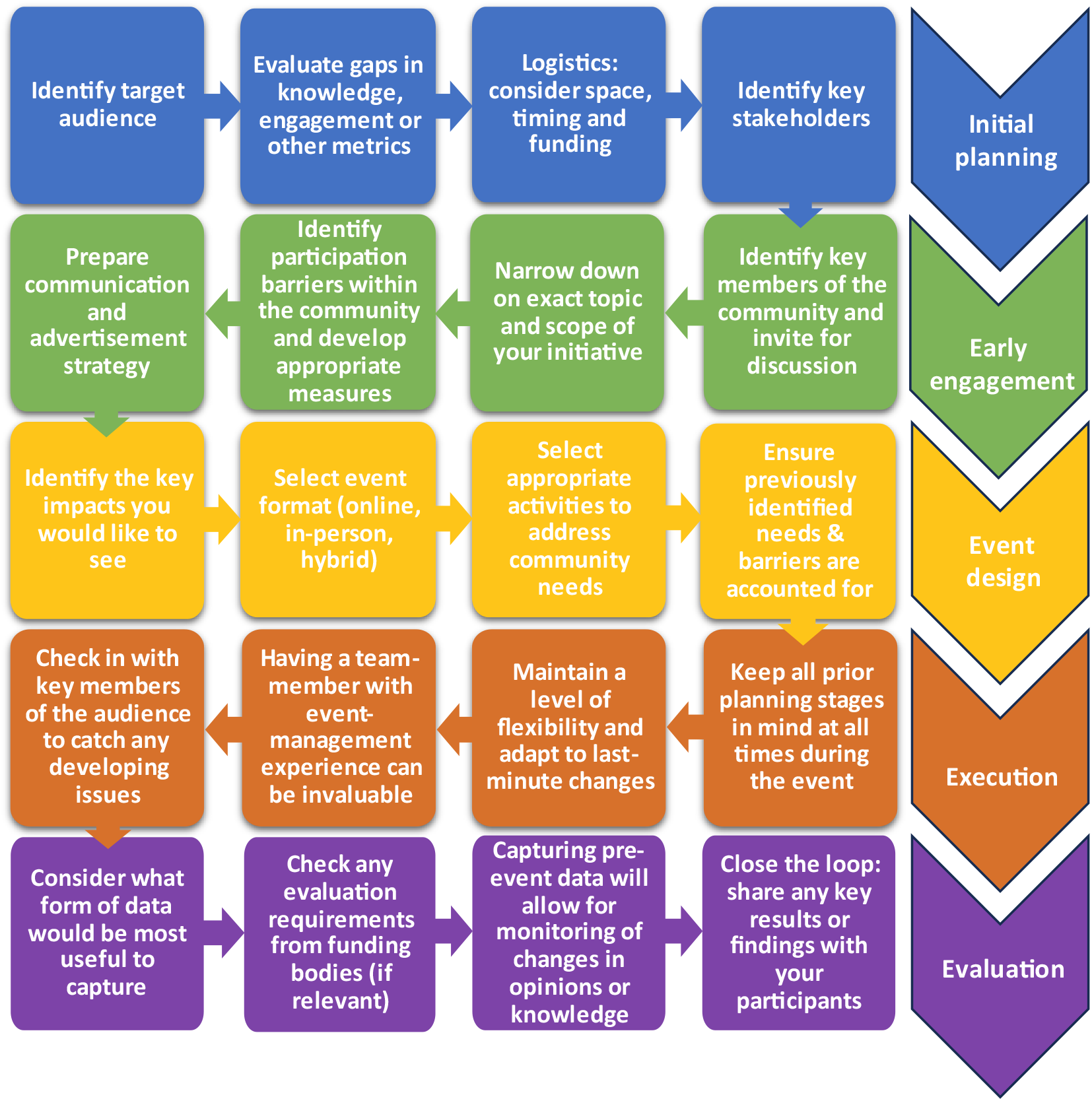
Organisational flow chart developed over four years of Invisible Spectrum initiatives.

### The Research Actors

This work required the collaboration of researchers and leaders within the target community. The engagement process began with a core-team of members of Precision Oncology Ireland (POI), a cancer research consortium of Irish universities, charities and industry partners, coordinated by Systems Biology Ireland (SBI; a research centre in University College Dublin). The POI team members included both academics and research support staff, all with a background in biological research. Crucially, we also engaged with scientists of Bangladeshi or Muslim background who were willing to help with the event and engage with members of the target community. This visibility of ethnic minority community members within the scientific community helped in building trust while also narrowing the cultural and language barriers between the two groups. One team-member had pre-existing links with the Bangladeshi community, which was utilised to establish and build relationships with this community.

The target audiences for our initial pilot initiative included both members of the Bangladeshi and Arabic Muslim communities in Ireland. However, this was later refined to just the Bangladeshi community in subsequent iterations. This group was selected based on the aforementioned barriers to research and healthcare access observed in this group. This is a small community: the most recent available census (2022) showed just 4,388 individuals originally born in Bangladesh now residing in the Irish state (12). Initial contact was first made with a small number of highly-engaged community ‘leaders’ to explain the vision of our programme, get early buy-in from the community and begin to establish a network of contacts. Once this group of community leaders was established, they were then invited to UCD to meet researchers and tour the laboratories, equipping them with the necessary knowledge to then act as ‘recruiters’ within the community and maximise reach to a wider audience.

### Co-design process

To co-design and deliver the programme, we worked in collaboration with the community leaders, with the aim of tailoring the programme to the needs and desires of the community and building a deeper trust with community members. Community leaders met with the researchers and visited the laboratories. Discussions held with community leaders were also used to shape recruitment strategy (e.g. social media community pages and WhatsApp groups popular within the community were identified and used for advertising the programme).

The specific contributions provided by community members included:

- Facilitating English-Bengali translation to create bilingual material.
- Providing important cultural context to help tailor aspects of our event to the community.
- Suggesting members of the community who could speak about their own experience with cancer.
- Suggesting appropriate and strategic avenues of community advertisement and promotion.
- Providing guidance on the knowledge level of the community to ensure programme content was developed at the appropriate level.
- Acting as “ambassadors” of the programme to the community, creating important links of trust and encouraging community members to attend and participate.
- Live-streaming of talks on the day to community-focused social media pages, to allow for virtual-attendance of those unavailable on the day.

We believe that the collaborative structure within our *Invisible Spectrum* programme is exceptionally strong and has allowed the programme to grow year-on-year. The co-design element of *Invisible Spectrum* is not a simple “box-checking” exercise but a real and crucial aspect of our organisational process, without which the programme would not have been a success.

### Data Collection and Analysis

Empirical data collection focused on the *Invisible Spectrum* events held over four years, with data including feedback polls conducted during the events and mixed-method pre- and post-event surveys. Polls were designed to capture participants’ experience of the events, including invitations to score different elements of the event. Surveys also included self-reported knowledge and confidence in discussing medical issues and satisfaction in the structure, delivery and content of events. Open-ended questions invited participants to make recommendations for the development and adaptation of the activities.

Quantitative data were analysed using GraphPad 8 (descriptive statistics and t-test) or SPSS 29 (ordinal regression, Wilcoxon signed-rank) software.

## Results

Invisible Spectrum has had four iterations, with changes to event structure between years based on feedback from attendees. Fig. 2 gives an overview of the evolving structure during this period. To capture this evolution, this section presents findings from each of the four years. As each annual iteration of the *Invisible Spectrum* programme varied in terms of thematic focus and format, we have organised this section to give an overview of each event from 2020 – 2023, with a final section focusing on findings from the 2022 event as an illustrative example of our data-gathering process.

**Figure 2.**
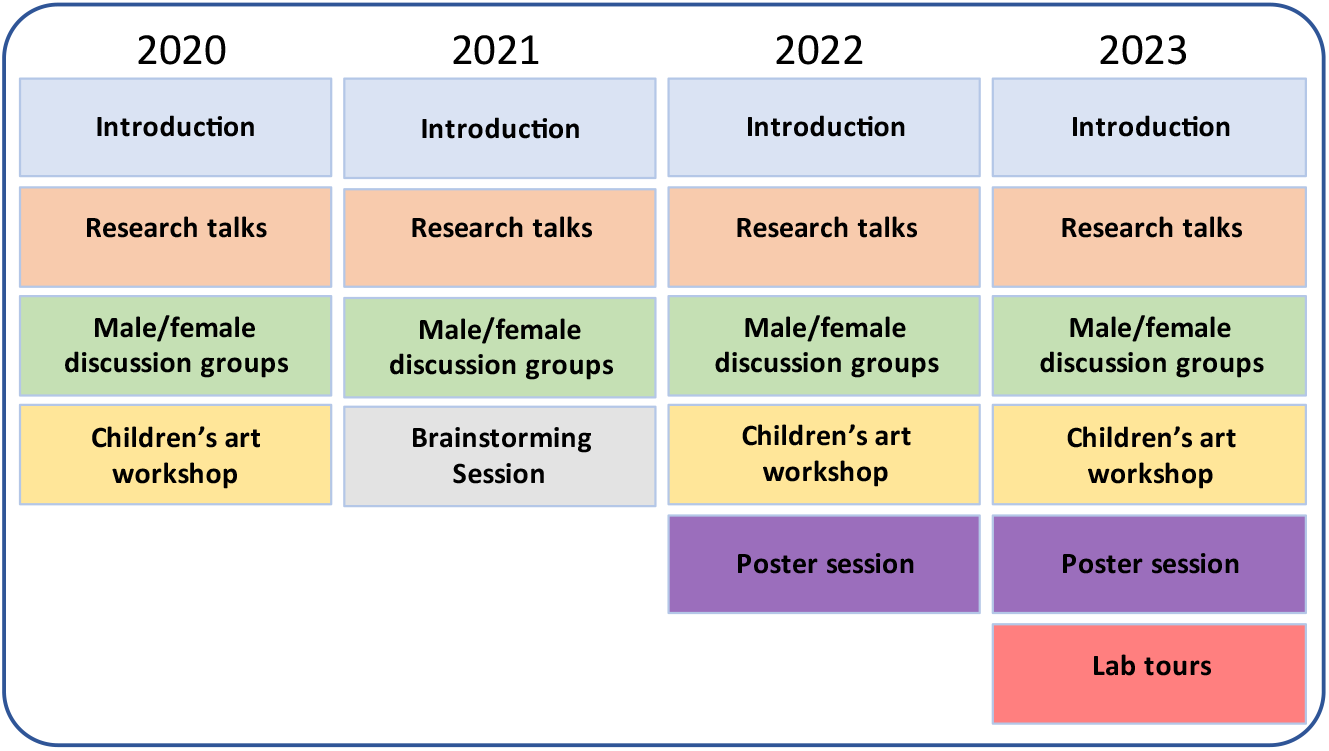
Event outline of Invisible Spectrum over four years

### 2020: Pilot event

The first *Invisible Spectrum* events were held in 2020, using an online format due to COVID restrictions. Separate events were held for each of the two target audiences: members of the Bangladeshi and Arabic Muslim communities. In total, we reached 106 participants across the two events (71 from the Bangladeshi and 35 from the Arabic Muslim communities). Each event had the same structure, consisting of a series of short interactive talks on cancer research and awareness, followed by a gender-segregated breakout discussion session, which were designed to alleviate some of the cultural sensitivities surrounding discussion of personal health-related issues. We also facilitated a children’s art competition.

Post-event feedback from the community provided some key recommendations, including suggestions to move to a face-to-face format, provide a mix of cancer research and cancer awareness content and allow additional time for discussion.

The key learnings from this first event were as follows:

- Our format was effective and well received by the community, especially the discussion sessions.
- The format had some room for improvement, such as lengthening the amount of time allocated for discussions.
- There was a desire within the community for more effective, face-to-face communication.
- The gender-segregated discussion sessions were well-received and encouraged a more open conversation about health-related issues.

### 2021: Increased prior-event engagement

After the success of our pilot programme, we refined several details for the 2021 iteration of *Invisible Spectrum*. In contrast to the previous year, we elected to focus our efforts solely on the Bangladeshi community. In order to increase programme efficacy and ensure audience needs were comprehensively addressed, we embarked on a major expansion of our community engagement efforts during the planning process, culminating in a series of one-hour interviews with 10 key members of the Bangladeshi community. These interviews covered a wide cross-section of community members, male and female, from a variety of backgrounds and social classes and included those living in Direct Provision Centres. The aim of these interviews was to probe the insights of the target community in terms of their engagement with science and research and explore the impact of socio-cultural factors on programme format and content.

Some of the key themes brought up during interviews included:

- The social stigma of discussing illnesses such as cancer, particularly among women
- The dual barriers of language and literacy in less-educated groups within the community
- “God will decide” - the prevalence of fatalism among the community
- “Bangladeshi women are shy” - the lack of confidence within community members to ask questions within a group, particularly a mixed gender group, and particularly women
- “Bangladeshi men are short of time” – men within the community are often time-poor as the primary financial provider in the family.
- “She will not get married” – the shunning of those with cancer within the community.
- “They are just hearing, they are not understanding or absorbing the information” – The crucial importance of social leaders in transmitting and translating the message.

Following the interview stage, we once again co-created an agenda that included research talks, gender-segregated discussions and post-event surveys. Due to ongoing COVID restrictions, we again opted for an online format, where we achieved similar attendance numbers as the previous year (approximately 130). We also created a more specific theme and branding than the previous year, choosing the topic of cancer genetics and “‘My DNA, My Health’ as a title.

Survey responses were collected before (n = 32) and after (n = 16) the 2021 event. Pre- and post-event surveys allowed for assessment of changes in perspectives and understanding. Attendees indicated increases in several measure of knowledge and confidence in discussing medical issues (Fig. 3A) as well as high levels of satisfaction in the structure, delivery and content (Figure 3B). We observed a 13% increase in attendee’s levels of confidence in discussing medical issues with their GP and a statistically significant 28% increase in attendee awareness of cancer screening programmes in Ireland (Fig. 3A, p<0.05). As with the previous year, feedback was also highly encouraging, with a majority of participants answering positively when queried about their perceived benefit from attending, their ease in following the content and their plans to attend future events (Fig. 3B).

**Figure 3.**
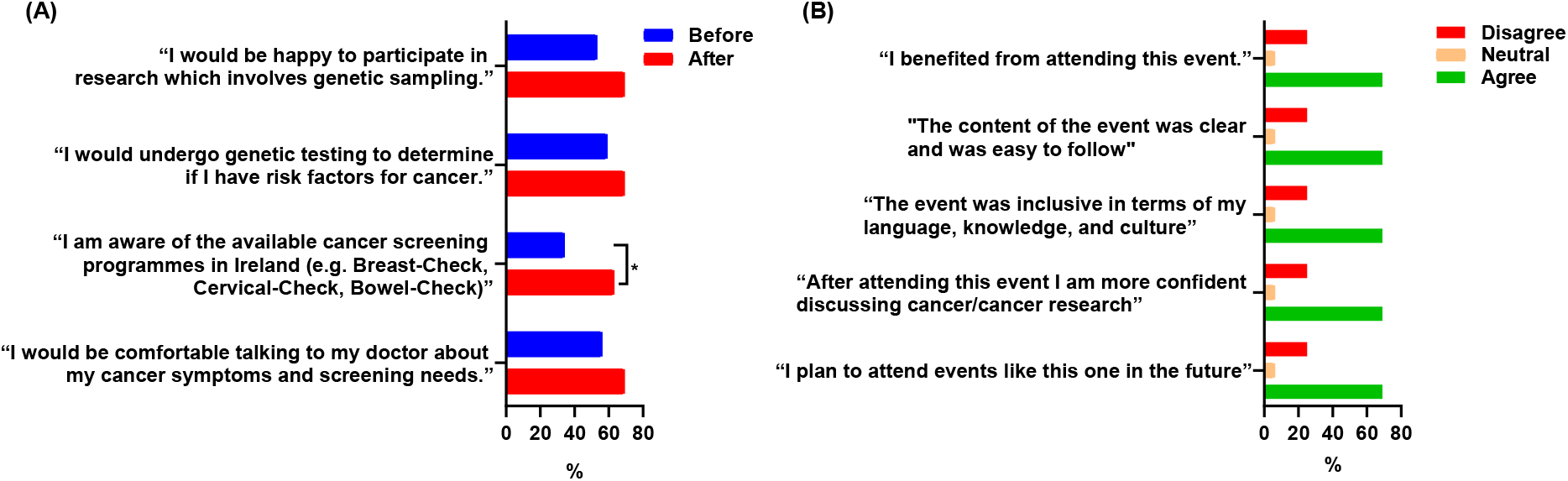
Highlight of survey results from 2021 Invisible Spectrum event.

### 2022: First in-person event

The lifting of many COVID restrictions in 2022 generated the opportunity to translate our successful event structure into an in-person format. This was a commonly requested format change from the community as it would allow for more interactive participation as well as the inclusion of young children in a separate engagement activity. Following a similar pattern to previous years, we again met with Bangladeshi community leaders to assess the needs of their community and worked together to co-design our first in-person event.

The 2022 *Invisible Spectrum* iteration was a bilingual family event under the theme of ‘Understanding Cancer’. The primary focus of the day was a series of talks, including an overview of basic cancer biology, screening programmes, cancer research programmes in Ireland and the importance of diverse inclusion in research. A display session of plain-English research posters gave participants the chance to meet and converse with young cancer researchers in a relaxed setting and ask further details about their work. Several of the presenting researchers and organisers were Bengali speakers which further assisted in building trust, removing barriers and promoting open communication.

Similar to previous years, we included a gender-segregated breakout session, where a representative from the Irish Cancer Society provided a brief overview of the diverse range of symptoms that may warrant further consultation with a medical professional. This part of the day was particularly impactful and there was a robust back and forth dialogue, stretching beyond the initially allocated time.

Finally, we incorporated an educational children’s activity, which consisted of a “Cell-building workshop” and “Pipette Art”, with the aim of encouraging scientific interest as well as allowing the parents freedom to focus on the talks and workshops.

Moving the *Invisible Spectrum* programme to an in-person format presented additional challenges (detailed below) and the input and assistance of community leaders was highly valuable in ensuring that community accessibility remained at the forefront of our organisational efforts.

- As in previous years, all talks were delivered bilingually in English and Bengali (requiring some additional planning to accommodate the difficulty of live translation).
- The event itself took place in a hotel function room, at a location that was deemed to be convenient and accessible by community members.
- A weekend date was selected to ensure the greatest level of community participation by reducing difficulties with participants work schedules while also allowing for participants to bring children.
- The event was followed by a sit-down lunch for the group, which allowed for casual interactions and follow-up discussions.

### 2023: Focused information

The most recent *Invisible Spectrum* event, held in November 2023, incorporated previously established programme segments (research presentations, poster sessions, bilingual discussion groups and children’s activities) along with two significant modifications. We opted to host the event within the SBI research institute itself, which allowed us to include tours of the research facilities. Facilitated by early-career researchers, these lab tours were extremely well received by the community and served a vital purpose of “demystifying” the process of research and giving them a sense of the general layout and workflow of a modern research facility, as well as witnessing some standard laboratory procedures. It also provided an excellent method of reinforcing points brought up in earlier presentations (e.g. providing samples of cells for attendees to view under a microscope and linking demonstrations of research techniques to previously discussed results). As an additional advantage, we also reserved a private lab-tour slot for attendees of secondary-school age and used the “introductory talk” part of the tours to emphasise the potential benefits of studying science within university, which helped fulfil our secondary ambitions of encouraging greater participation of the Bangladeshi community within science and academia.

We also created a more focused structure within the gender-segregated breakout sessions. In contrast to the broad discussions on general cancer symptoms and screening utilised in previous events, we instead elected to focus each session on a particular form of cancer of significant relevance to that group, with the male room concentrating on colorectal cancer and the women’s session looking at ovarian cancer. These cancer types were selected following discussion with Bangladeshi community leaders and are of significant clinical relevance to Bangladeshi migrant communities: studies from the UK have found low levels of uptake of colorectal screening programmes in Bangladeshi men (13) and low levels of awareness of ovarian cancer symptoms in South-Asian (including Bangladeshi) women (14).

Each group participated in a focused discussion on symptoms, risk-factors and proactive preventative measures on their respective cancer type, with information presented bilingually. The goal of this change was to build on prior engagement with the community and leverage their increased understanding of cancer in order to provide a more targeted informational benefit within selected cancer types of relevance to their community. When scored on their ability to recognise potential symptoms of colorectal (male group) or ovarian cancer (female group) in paired pre-and post-event surveys, we observed enhanced recognition in the male group (Fig. 4A, 52% v. 81%, n = 12, p<0.05). In the female group, there was a strong trend towards increased recognition but this did not reach statistical significance, possibly due to a lower sample size (Fig. 4B, 63% v. 96%, n = 6, p = 0.12).

**Figure 4.**
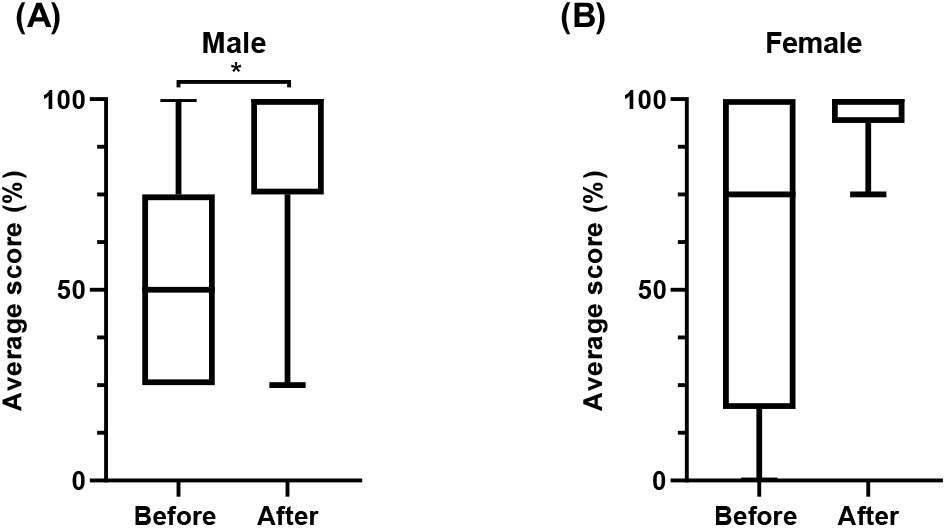
Awareness of cancer symptoms in male and female Invisible Spectrum 2023 participants.

### 2022: Detailed reporting

Evaluation and gathering of feedback from each iteration of *Invisible Spectrum* has allowed for a continual refinement of the programme structure. As a more detailed example of the process, we have included an analysis of qualitative and quantitative data (Figs. 5 & 6) from the 2022 programme. This was accomplished via matched pre-event and post-event surveys as well as open feedback invitations.

**Figure 5.**
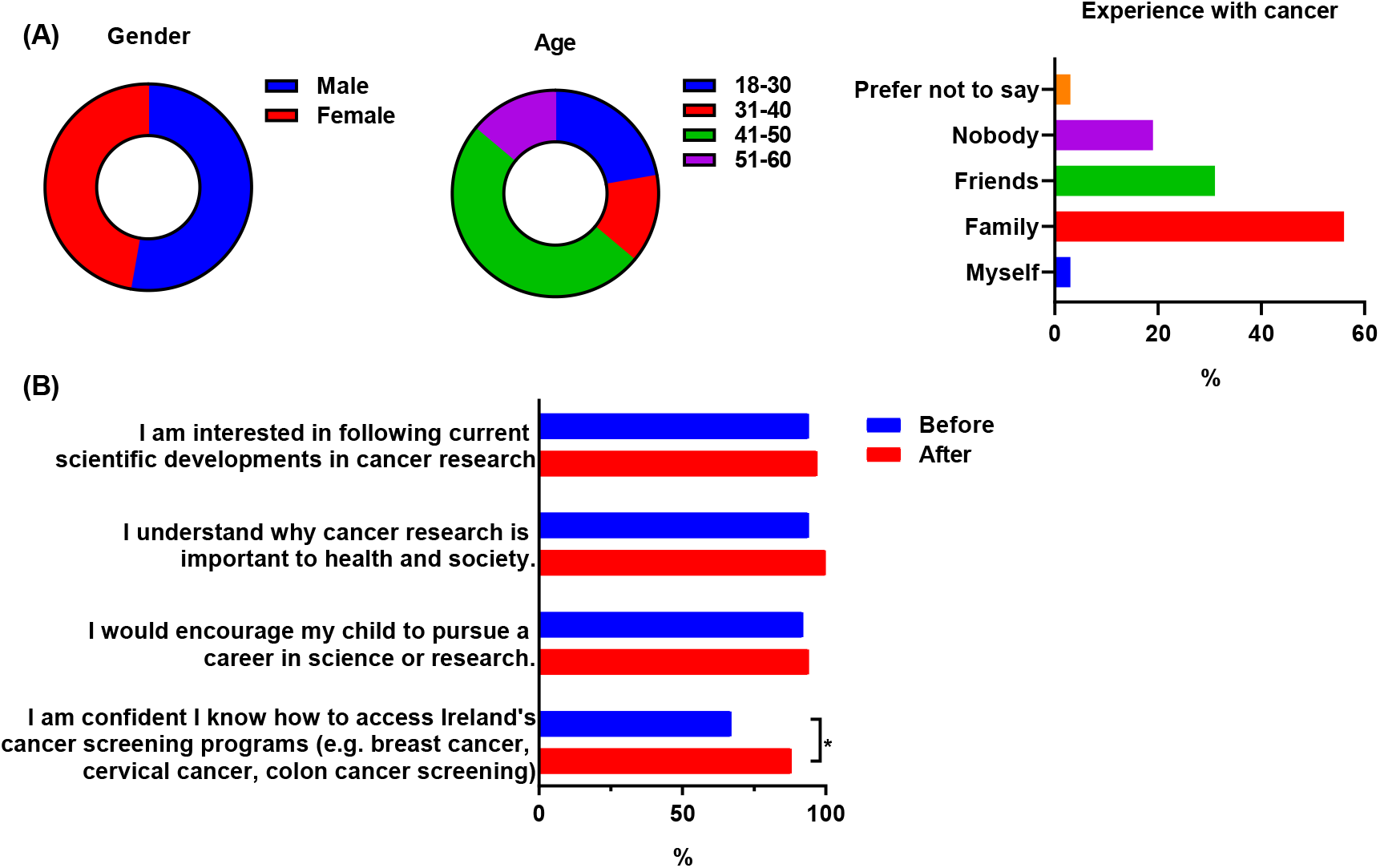
Breakdown of participant demographics and baseline interest in science

Attendees answered basic demographic questions regarding gender, age and experience with cancer in a pre-event survey. Our participants (n = 36) were evenly distributed between male and female (47% v. 53%, Fig. 5A), with the majority (64%) being aged between 40 and 60 years. These age brackets constitute a key demographic for promotion of cancer screening and healthcare intervention.

When surveyed regarding their own personal experiences with cancer, there was widespread experience of having someone close to them affected; with 56% having a family member diagnosed with cancer, 31% with a friend and only 18% responding with no close personal experience (Fig. 5A). We had just one survey response from an individual with a cancer diagnosis. Finally, data gathered to measure levels of interest in research indicated that attendees had high levels of interest and engagement (for example, 94% would encourage their child to pursue a career in science and 100% agreed they understood the importance of cancer research, Fig. 5B). Of interest, data from the same question in a post-event survey showed a significant increase in confidence in accessing cancer screening services (66% vs. 88%, p<0.05).

A “True/False” questionnaire in pre and post surveys was used to measure two key outcomes: understanding of cancer biology (assessed with questions surrounding cancer genetics, screening and basic biological concepts) and as a secondary outcome, participant recognition of the benefits of a precision medicine approach to cancer therapy as well as awareness of the POI programme itself.

Fig. 6A shows the percentage of attendees answering each question correctly in both pre and post surveys. The main questions that we observed improved scores on were “Understanding a person’s genes and proteins can allow doctors to personalise treatment” and “The POI programme is the largest cancer research programme currently in Ireland”. In total, 40.5% of participants improved their scores over the course of the event, indicating an improved average score across all participants of 3.9 to 4.6 when comparing pre and post-event surveys (Fig. 6B, p<0.05).

**Figure 6.**
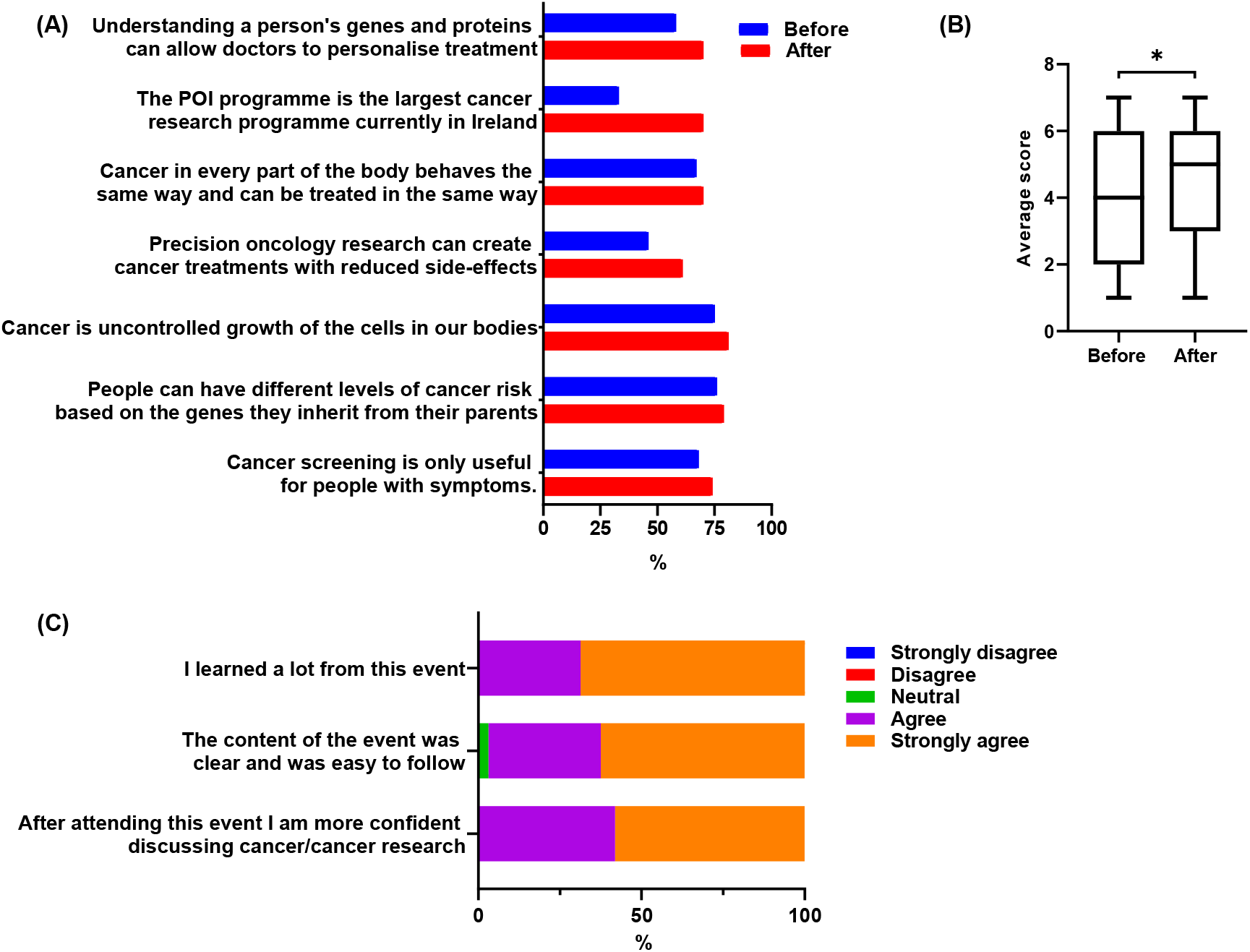
Evaluating learning outcomes

Finally, post-event feedback indicated a high level of satisfaction with event content and delivery, with near universal agreement with the statements “I learned a lot from this event”, “The content of the event was clear and was easy to follow” and perhaps most importantly, “After attending this event I am more confident discussing cancer/cancer research” (Fig. 6C).

### Final reflection

In addition to quantitative data, we also gathered qualitative data and feedback each year, both via the surveys described above but also via informal routes including discussions with attendees after each event and emails received to the team. Although we received many positive and highly encouraging quotes, one quote, shown below, is worth highlighting as it encapsulates many of the difficulties and challenges faced by overlooked communities and how targeted engagement initiatives, like the one described here, can be effective tools for remedying these problems.

> *“Being a cancer survivor myself, I can not stress enough the importance of early screening of cancer. In my community, the word “cancer” is still a taboo and people fear it. The more we will know about it, the less we will be afraid of it. Thanks to all for organising the event where people of my community could get the knowledge and know about the screening opportunities. This will surely help a lot of us. “*

It should also be noted that this quote came from a member of the community who has been closely involved with the *Invisible Spectrum* programme over a number of years, providing assistance with event planning, translating and promotion. Having these kinds of individuals as part of our programme is of crucial importance for continually growing our programme and strengthening ties with the engaged community.

## Discussion

The *Invisible Spectrum* programme was designed from the ground up, with community support, to address a real need within ethnic-minority communities for increased engagement and targeted information around cancer awareness. Results from other groups highlight the need for these kinds of interventions. A study of 37 ethnic minority women in Denmark, using a similar structured interview format to what we deployed in our early planning stages, found a range of reasons preventing them from further engagement with cancer screening services (15). These ranged from language barriers, socio-cultural (particularly a discomfort with physical examination amongst female members of the community), inadequate understanding of the role of cancer screening and prevention (e.g., a belief that only those with symptoms need to be screened) and a distrust in the Danish medical system. Many of these concerns were also raised in our cohort, suggesting these are common barriers applicable to ethnic-minority communities in many different countries.

Importantly, these studies highlight not only barriers that need to be challenged but also new potential avenues by which existing efforts can be adapted to better efficacy. A strong reliance on oral, inter-group communication rather than written text is a common finding in minority communities (15, 16). This is something we also identified early in our programme and has led to us deploying a strong focus on oral communication networks and “word of mouth” when promoting each year’s event.

Ongoing communication and partnership with figures from the Bangladeshi community in Ireland allows us to identify existing barriers faced by the community limiting participation in cancer screening and other vital medical services, including language and cultural barriers, lack of understanding of cancer symptoms and treatment options, misconceptions such as the ineffectiveness of screening programmes in symptomless people and discomfort with open discussion of personal medical issues.

We are continually developing and improving our practices and procedures to draw in wider cohorts from the community and produce the maximum possible impact. Each year, we implement measures to ensure that any possible barriers to entry are reduced, if not eliminated. These measures have included:

- Provision of bilingual material and content. This includes not only all advertising flyers, surveys and other written communications but also a live-translation process for all talks and Q&A sessions.
- Careful consideration of the cultural background of the community and how it can be accommodated. A major example of how this was enacted was creating gender-separated breakout sessions, with male or female researchers/clinicians present in each separate area to allow for frank and open discussion that would not have been possible without this provision. When events have been held in the evening, we have also provided space for participants to perform their evening prayers when necessary.
- Understanding the family-focused nature of the community and, where possible, facilitating young children as part of the event. We created a highly interactive, science-based arts workshop that ran in parallel with the talks and discussions, both keeping the younger members of the community occupied and also creating a secondary learning environment This further highlights the essential nature of involving community members right from the start as they have the best chance of motivating and recruiting less engaged members of their community to participate in these events.

Before concluding, it is important to reflect on the process of evaluation captured in this paper. Meaningful engagement with the community was a central focus of our work, but the ability to gather qualitative and quantitative data in the course of our activities provided an important source of guidance for the team. At the same time, while recognising the small samples completing data collection invitations as part of the events, the data provide an important illustration of participants’ experiences of these activities. They were an important part of the evolution of the programme, but also provided an initial assessment of the potential impact and benefits of this type of activity. However, the analyses completed cannot be taken as a comprehensive assessment of the programme.

In conclusion, over the course of 4 years of the *Invisible Spectrum* programme, we have developed and implemented an effective organisational structure, built on regular community feedback. We believe this to be an effective model of community engagement and hope that some of the learnings contained within this article can be successfully adapted and utilised by others wishing to host similar initiatives.

## Data Availability

All data produced in the present study are available upon reasonable request to the authors

## Declarations

### Ethics Approval

This study received “low-risk” ethics approval from University College Dublin’s Human Research Ethics Committee – Sciences (HREC-LS).

### Funding

This publication has emanated from research conducted with the financial support of Precision Oncology Ireland, a Consortium of 5 Irish Universities, 6 Irish Charities, and 8 Industry Partners, which is part-funded by the Taighde Éireann – Research Ireland Strategic Partnership Programme, under Grant number [18/SPP/3522].

### Competing Interests

The authors declare no competing interests.

### Author Contributions

WK, AR, WMG, JCR, EMK & FL conceptualized the initial pilot study. WK, AR, WMG, JCR, EMK, FL, TM, SS & SOG co-organised subsequent initiatives. WK, JCR, EMK & SOG developed the manuscript’s strategic objectives and structure. SOG performed data collection, data analysis and drafted the manuscript. All authors gave critical insights and strategic direction to the initiative. All authors read, provided feedback, and approved the final manuscript.

## Notes

### Competing Interest Statement

The authors have declared no competing interest.

### Author Declarations

Human Research Ethics Committee Sciences (HREC-LS) of University College Dublin gave ethical approval for this work.

